# Prospective Trial Registration and Publication Rates of Randomized Clinical Trials in Digital Health: A Cross Sectional Analysis of Global Trial Registries

**DOI:** 10.1101/19004390

**Authors:** Mustafa Al-Durra, Robert P. Nolan, Emily Seto, Joseph A. Cafazzo

**Author notes:** Corresponding Author: Mustafa Al-Durra, BSc MSc, Institute of Health Policy, Management and Evaluation, The Dalla Lana School of Public Health, University of Toronto and the Centre for Global eHealth Innovation, Techna Institute, University Health Network. Toronto General Hospital, R Fraser Elliott Building, 4th Floor 190 Elizabeth Street, Toronto, ON, M5G 2C4, Canada.

## Abstract

Registration of clinical trials was introduced to mitigate the risk of publication and selective reporting bias in the realm of clinical research. The prevalence of publication and selective reporting bias in trial results has been evidenced through scientific research. This bias may compromise the ethical and methodological conduct in the design, implementation and dissemination of evidence-based healthcare interventions. Principal investigators of digital health trials may be overwhelmed with challenges that are unique to digital health research, such as the usability of the intervention under test, participant recruitment, and retention challenges that may contribute to non-publication rate and prospective trial registration. Our primary research objective was to examine the prevalence of prospective registration and publication rates in digital health trials. We included 417 trials that enrolled participants in 2012 and were registered in any of the seventeen WHO registries. The prospective registration and publication rates were at (38.4%) and (65.5%) respectively. We identified a statistically significant (*P*<.001) “Selective Registration Bias” with 95.7% of trials published within a year after registration, were registered retrospectively. We reported a statistically significant relationship (*P*=.003) between prospective registration and funding sources, with industry-funded trials having the lowest compliance with prospective registration at (14.3%). The lowest non-publication rates were in the Middle East (26.7%) and Europe (28%), and the highest were in Asia (56.5%) and the U.S. (42.5%). We found statistically significant differences (*P*<.001) between trial location and funding sources with the highest percentage of industry funded trials in Asia (17.3%) and the U.S. (3.3%).

## Introduction

In the realm of scientific research, study results should be made available and accessible to the broader research community to assess the evidence around the efficacy and potential harm of healthcare interventions.[1] Emanuel et al.’s framework defines seven ethical guidelines for clinical research involving human subjects.[2] The first of which describes the value derived from disseminating research results as *“Only if society will gain knowledge, which requires sharing results, whether positive or negative, can exposing human subjects to risk in clinical research be justified”*.

A number of studies have examined the ramification of publication bias and debated a number of contributing factors to non-publication of clinical trial results such as recruitment challenges, funding sources and study design.[3-10] Other challenges pertaining to investigators were also identified as contributing factors to non-publication of clinical trial results, such as lack of time or disagreement with coauthors.[11,12]

The notion of clinical trial registration was conceived in 1986, when Simes suggested that the act of registration would help control the risk of publication bias by providing a new data source for trial information and results.[13-15] The trial registries would also help mitigate selective reporting of positive outcome by comparing outcomes reported in trial publication versus outcome measures indicated in the trial registration.[16-21] In 2004, the International Committee of Medical Journal Editors (ICMJE) introduced a new mandate to promote prompt registration of all clinical trials.[22] In 2005, the World Health Organization (WHO) started an initiative to standardize trial registrations and trial registry datasets across multiple national and international registries.[23] In October 2008, the 7th revision of the Declaration of Helsinki was adopted with new requirements focusing on the importance of prospective registration of clinical trials and reporting of their results.[24] The prospective registration of clinical trial means that investigators should register their trials prior to the enrollment date of the first trial participant, otherwise, the registration would be considered retrospective. In 2015, the WHO announced a new statement on public disclosure of clinical trial results with more guidelines on trial registration, publication of results, and the inclusion of the trial registration number (TRN) in respective publications to enable linking of trial reports with clinical trial registry information.[25] As of September 2^nd^, 2018, the WHO International Clinical Trials Registry Platform (ICTRP) includes seventeen different national and international trial registries with a unified search and access to registration information of 441,033 unique clinical trials.[26]

Two studies reported publication rates between 66% and 68% for clinical trials registered in the US-based clinical trials registry, ClinicalTrials.gov.[27,29] Another study reported publication rates at 73% for clinical trials registered in at least one of several clinical trial registries (ClinicalTrials.gov, Current Controlled Trials, WHO International Clinical Trials Registry Platform, Clinical Study Register, and Indian, Australian-New Zealand, and Chinese Clinical Trial Registries).[28] Three other studies investigated prospective trial registration and the quality of registration in the International Standard Randomized Controlled Trial Number (ISRCTN) registry and the WHO ICTRP registries.[30-32] The results of these studies reported that prospective trial registration was between 37.8% and 53.4%.

However, investigators could be overwhelmed with challenges that may be particularly problematic in digital health trials, such as the usability of the intervention under test, participant recruitment, and retention challenges that may contribute to non-publication rate and prospective trial registration.[33-39] To our knowledge, this is the first review to analyze the non-publication rate and prospective registration of digital health clinical trials. We sought to examine, at the global level, the non-publication and prospective trial registration rates in digital health trials across the seventeen WHO ICTRP registries.

### Research Objectives

The primary research objective was to examine the prospective trial registration and publication rates of digital health randomized clinical trials registered in any registry that is part of the WHO ICTRP registries. The secondary research objectives were (1) to investigate the compliance with recruitment and inclusion of the TRN in the published trials, (2) to explore the relationship between publication rates, the time to publication and trial size, and (3) to analyze the relationship between retrospective trial registration and the duration from trial registration to the publication of trial results.

## Results

A full search of the WHO ICTRP database returned 441,033 unique clinical trials (507,455 in total including 66,442 duplicates) as of September 2nd, 2018. We utilized the advanced search functionality on the ICTRP search portal to apply our 86 search terms and phrases including all trial phases and recruitment status. There were 22,859 unique trials that matched our search terms, within which, 15,096 trials were randomized, and 1,018 trials were enrolled in 2012. After screening against our inclusion and exclusion criteria, 417 trials were included, and 601 trials were excluded as per the following breakdown:

### Prospective Trial Registration and Publication Rates

In summary, 273/417 (65.5%) trials were associated with identified outcome publications and 144/556 (34.5%) trials did not have any identified publications as shown in Figure 2:

**Figure 1.**
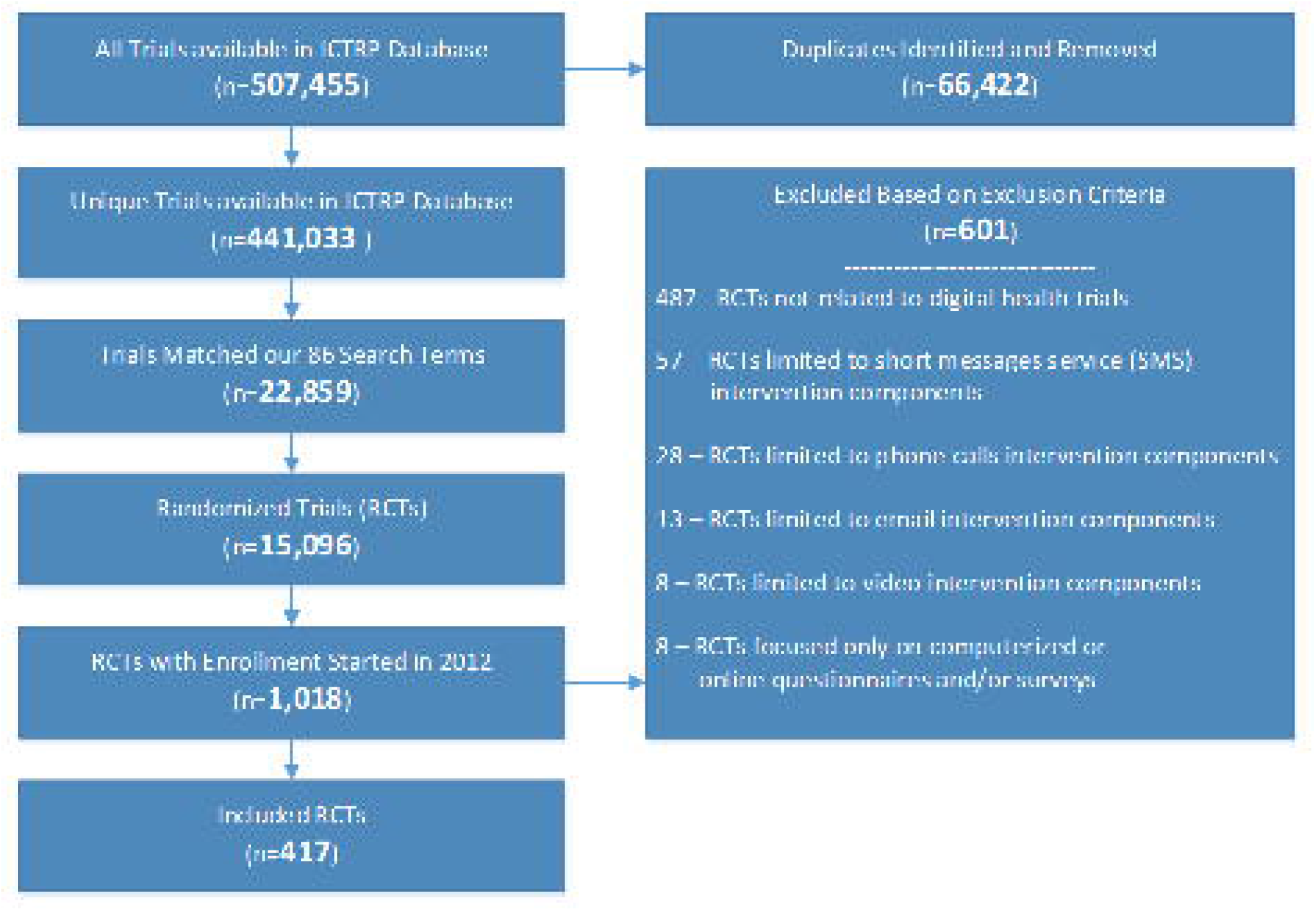
Included Trials from the Search Results

**Figure 2.**
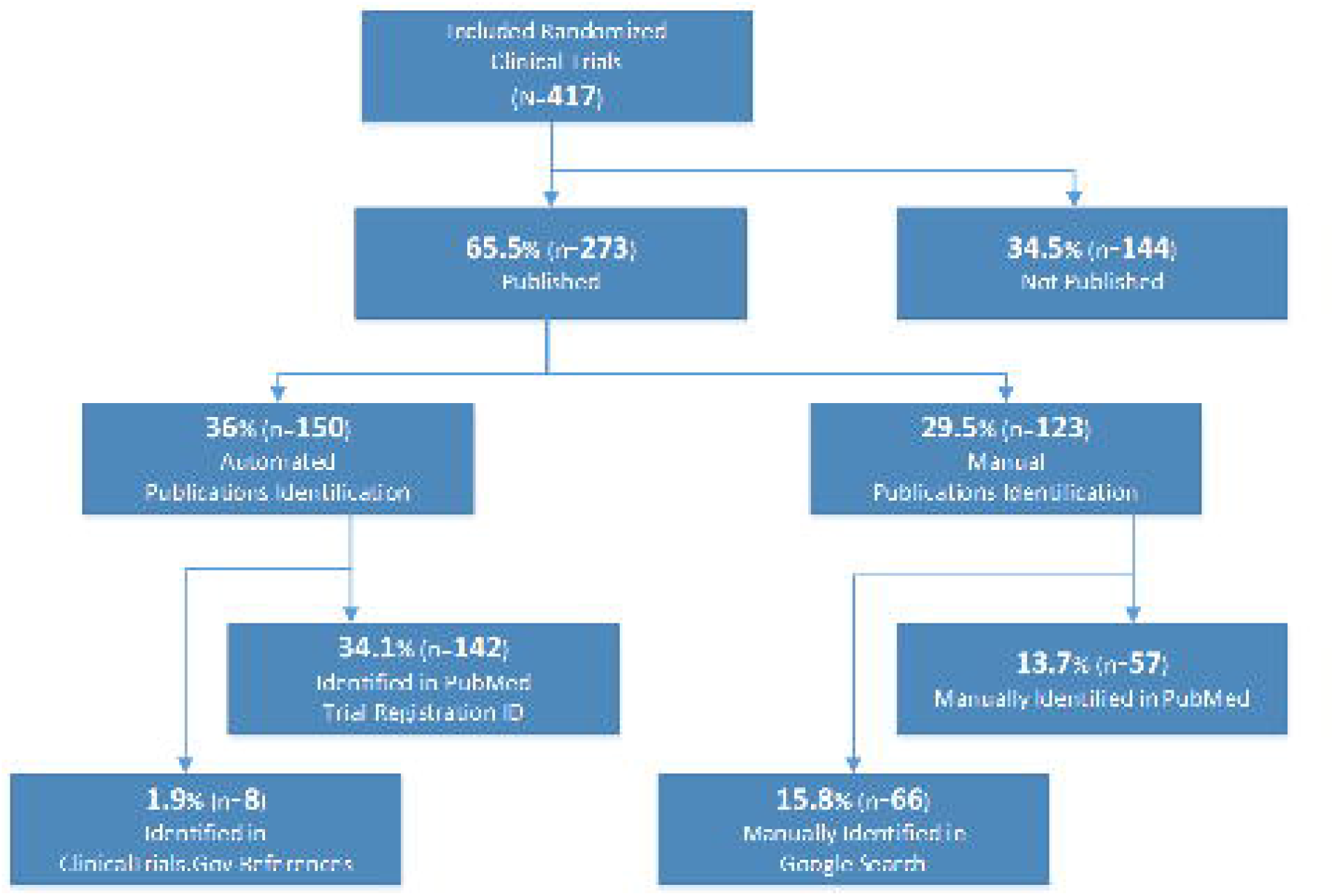
Publication Identification Results

We examined the relationship between trial characteristics and the non-publication rate and prospective registration of all included trials as shown in Table 1:

**Table 1.**
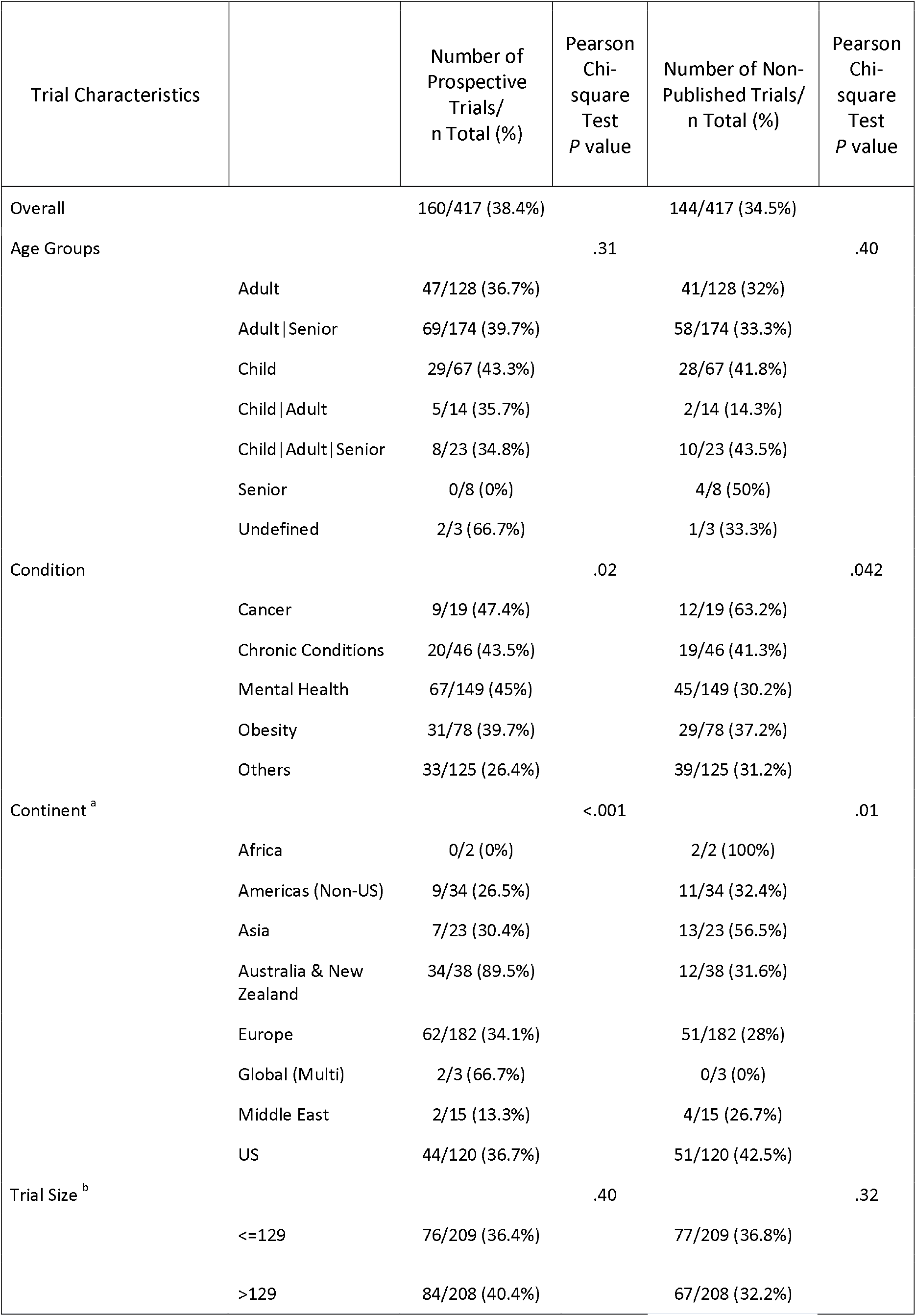
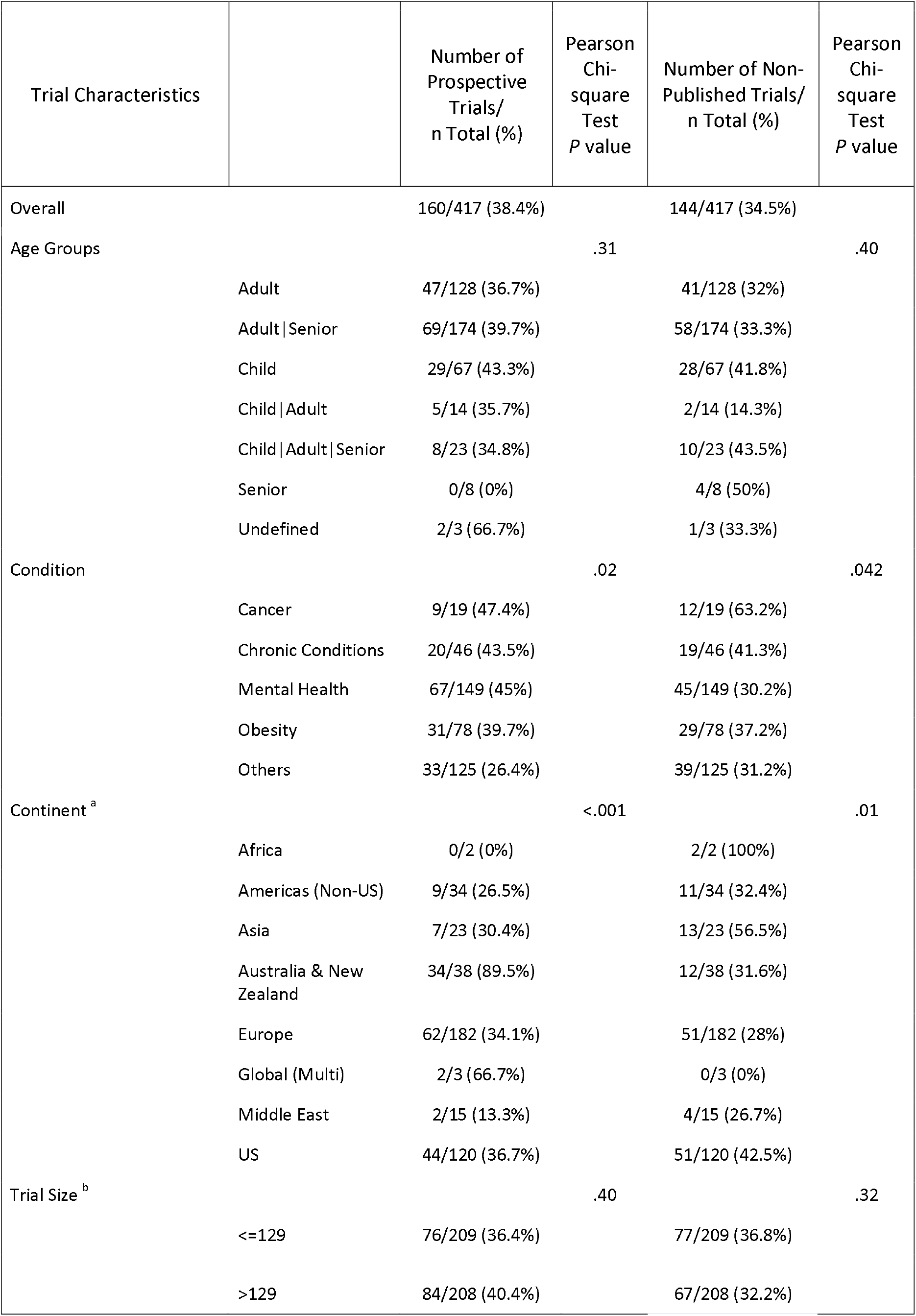
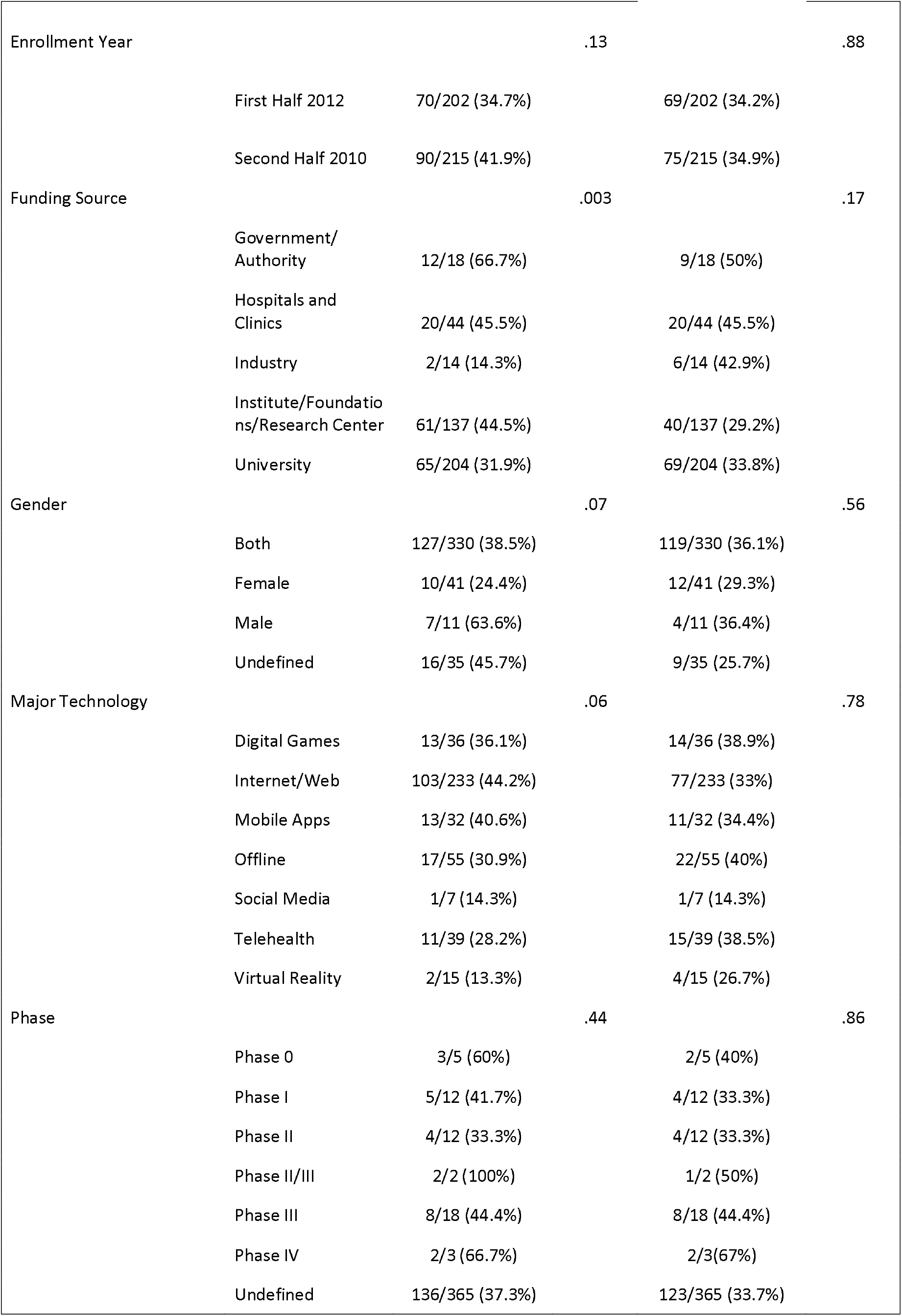
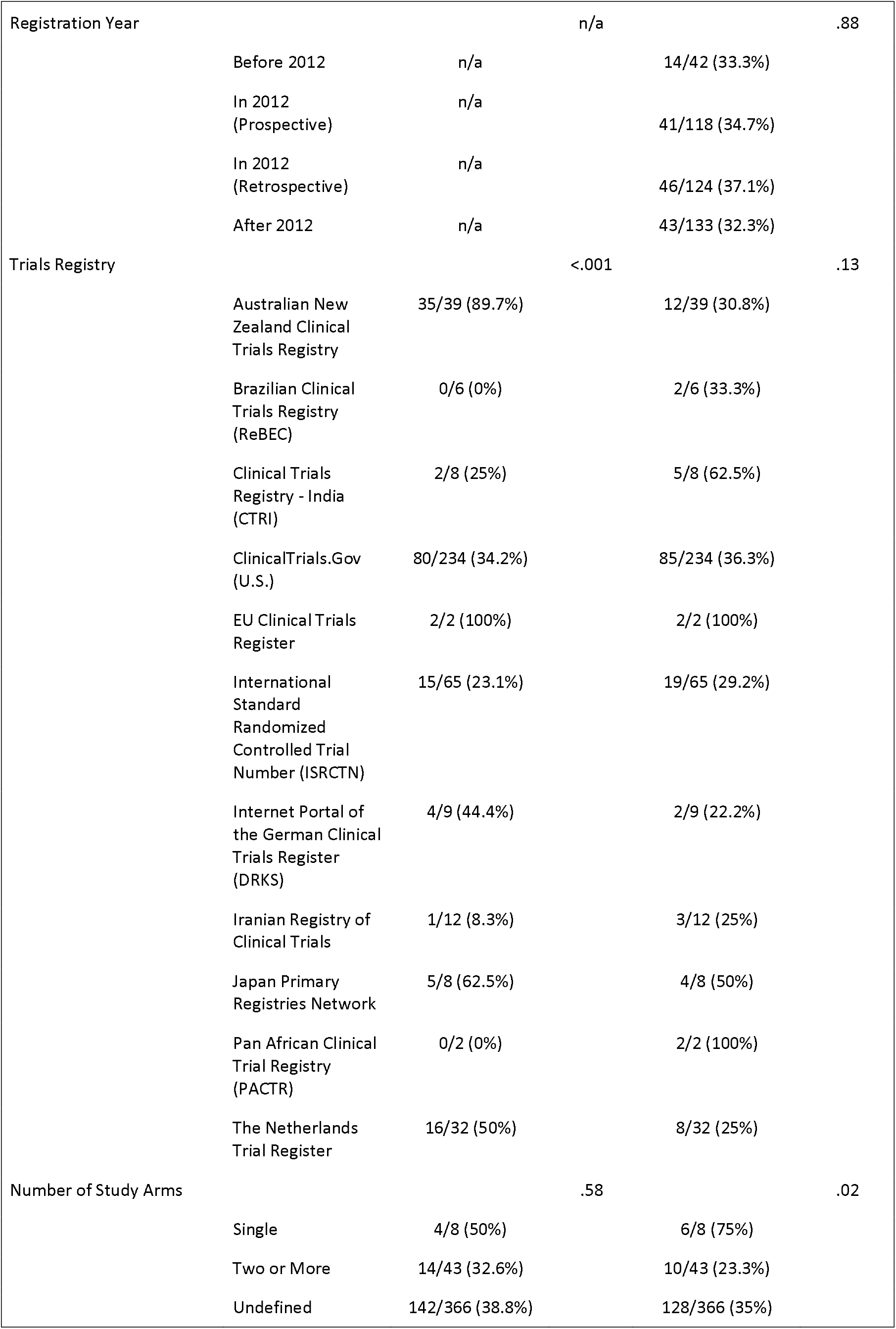
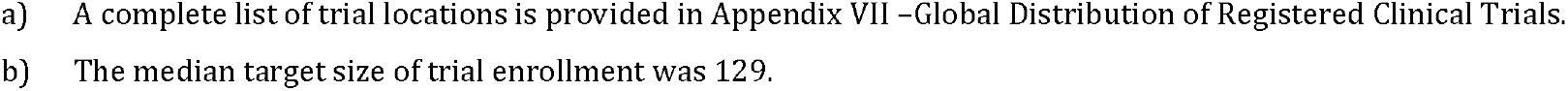
Relationship between Trial Characteristics and Prospective Registration and Non-Publication Rates of Included Trials

The Pearson Chi-square test results found significant relationships (*P*<0.05) between trial non-publication rate and trial characteristics including trial conditions, locations, and number of study arms. There were no significant relationships between trial non-publication rate and trial age group, enrollment date, funding source, gender, major technology, registration date, trial phase, trial registry, and whether the trials were registered prospectively.

Only 160(38.4%) of the included trials were registered prospectively. There were significant relationships (*P*<0.05) between prospective trial registration and trial characteristics, including trial conditions, funding source, location, and trial registry.

Our results showed that 144(34.5%) of the included trials remain unpublished. There were significant relationships (*P*<0.05) between the non-publication rates and trial characteristics, including trial’s condition, location, and study arms.

The highest non-publication rates were reported in Asia and the U.S. at 56.5% and 42.5% respectively. We did not consider the 100% non-publication rate of digital health trials in Africa due to the limited number of two trials included in our study. The highest percentage of industry funded trials was reported in Asia and the U.S. at 17.3% and 3.3% respectively. To interpret these results, we examined the relationship, and found statistically significant differences (*P*<.001), between the trial location and funding sources as indicted in Appendix VI – Funding Sources by Trial Registry and Location.

### Enrollment-To-Publication Duration and Trial Size

We postulated that smaller trials would be easier to conduct as they have less recruitment and enrollment challenges, hence they would likely be completed and published in a shorter time compared to larger trials. To validate our postulate, we analyzed the relationship, and found statistically significant differences (*P*=.002), between the trial size and the enrollment-to-publication duration. The enrollment- to-publication time was measured as the duration between the enrollment date and the publication date of the included and published trials, details provided in Appendix VIII – Relationship between Trial Enrollment-To-Publication Duration and Trial Size. We also explored the trend between enrollment-to-publication duration, trial size, number of published trials and the cumulative percentage of non-publication rate of all published trials. Details can be found in Appendix IX – Summary of Trial Enrollment-To-Publication Duration, Number of Published Trials, and the Cumulative Percentage of Non-Publication Rates. The results are depicted in Figure 3 revealing an incremental trend of published trial until a critical time point at the 4th year, where fewer trials were published into the 5th and 6th year after enrollment.

**Figure 3.**
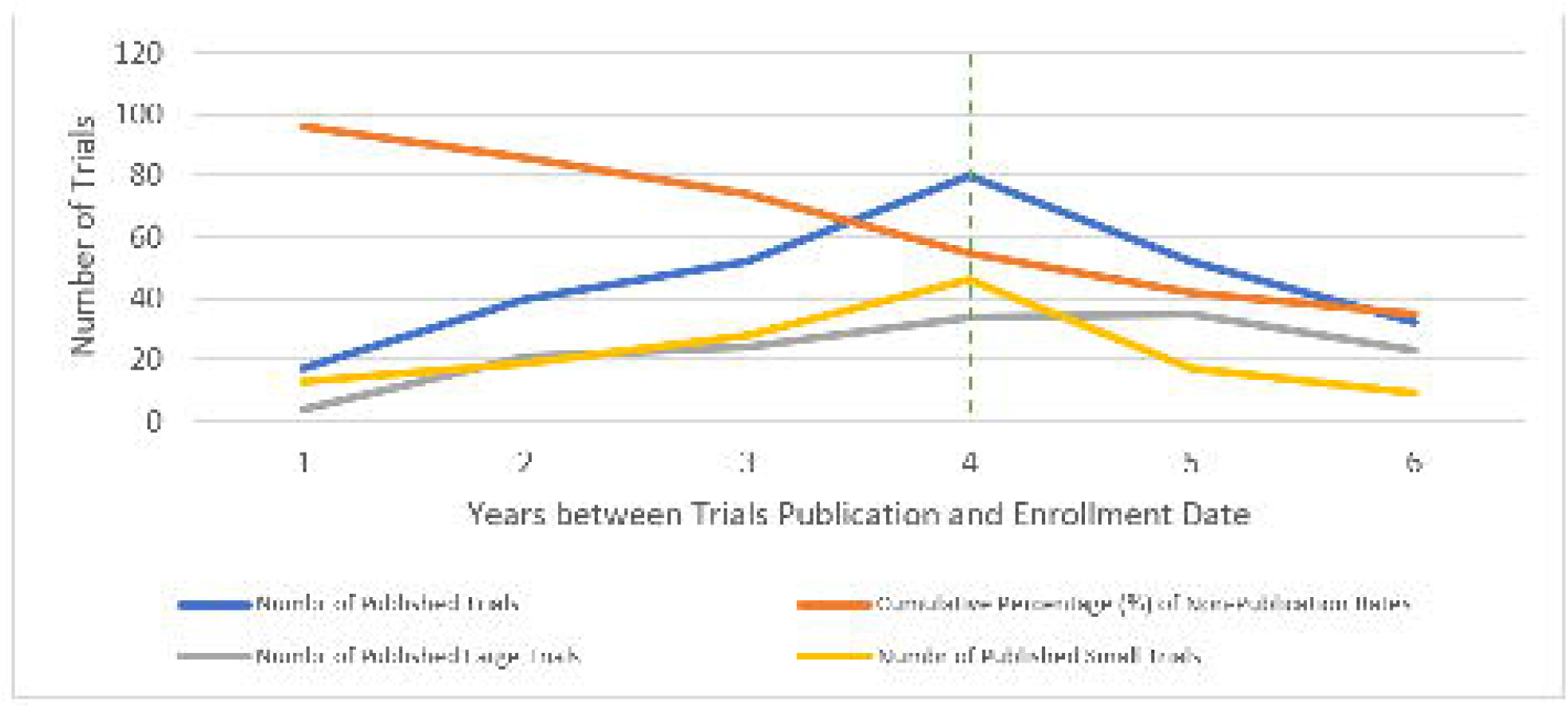
Relationship between Trial Enrollment-To-Publication Duration and Trial Size, Number of Published Trials and Non-Publication Rate

### Registration-To-Publication Duration and Retrospective Registration

We analyzed the registration-to-publication time as the duration between the registration date and the publication date of the included and published trials. We examined the prevalence of retrospective trial registration and its relationship to the registration-to-publication duration. We found a statistically significant relationship (*P*<.001) between retrospective trial registration and the registration-to-publication duration. The vast majority 95.7% of digital health clinical trials that were published within a year after trial registration, were registered retrospectively as shown in Table 4.

**Table 4.** Relationship between Trial Registration-To-Publication Duration and Retrospective Trial Registration

| Registration to Publication (Years) | Number of Retrospectively Registered Trials/ Number of Published Trials (%)<sup>a</sup> | Pearson Chi-square Test P value |
| --- | --- | --- |
| 1 or less year | 44/46 (95.7%) | <.001 |
| 2 | 30/48 (62.5%) |  |
| 3 | 34/52 (65.4%) |  |
| 4 | 30/57 (52.6%) |  |
| 5 | 21/45 (46.7%) |  |
| 6 | 9/22 (40.9%) |  |
| 7 | 0/3 (0%) |  |

a) Non-cumulative values. The number of retrospectively registered trials, the number of published trials, and the reported percentage is not cumulative within every year level.

#### Recruitment Compliance and Inclusion of the TRN in Published Trials

We sought to analyze the recruitment compliance in the 273 published trials. We compared the target trial size indicated in the registration information of the respective trials with their actual recruitment as reported in the identified publications. We found that 111(40.7%) of the published trials reported fewer subjects who were actually recruited than the target size indicated in the trial registry, 96(35.2%) published trials reported actual recruitment matching the trial target size, and 66(24.2%) published trial recruited more participants (over-recruitment) than the target size in the trial registry. Ensuring adequate participant recruitment is critical to support the statistical power and internal validity of the trial results. Enrollment of fewer participants could introduce type II errors in reported study results.[81] We suggest that over-recruitment could be appropriate and would empower further assessment of secondary hypotheses. Therefore, we considered a trial recruitment as compliant if the actual recruitment was equal to or more than the target size as defined in the trial registration.

We also analyzed the inclusion of the TRN in published trials. We verified whether a reference to the TRN was indicated in the papers of the 273 published trials. We examined the relationship between trial characteristics and the recruitment compliance and inclusion of the TRN in published trials as shown in Table 5.

**Table 5.**
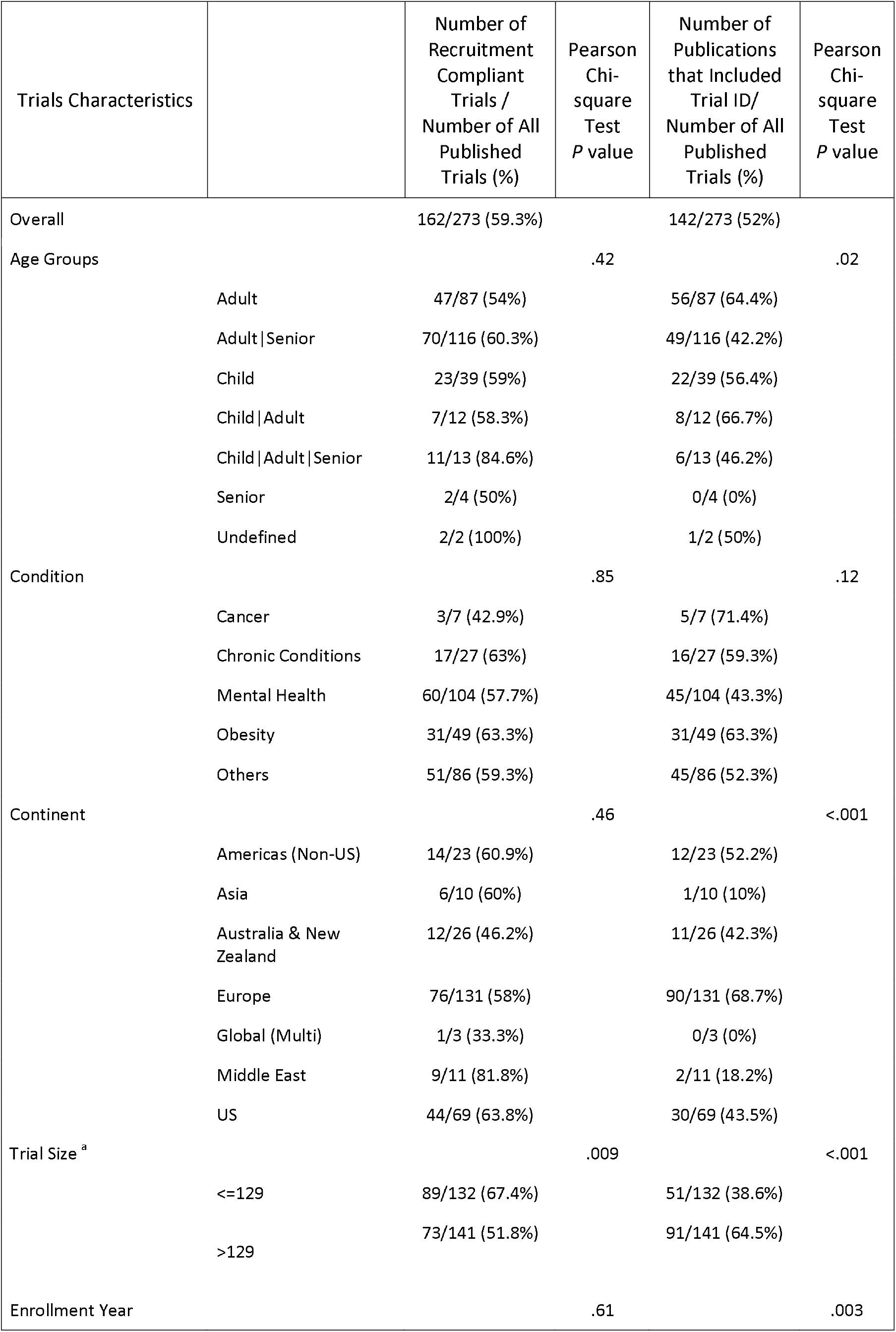
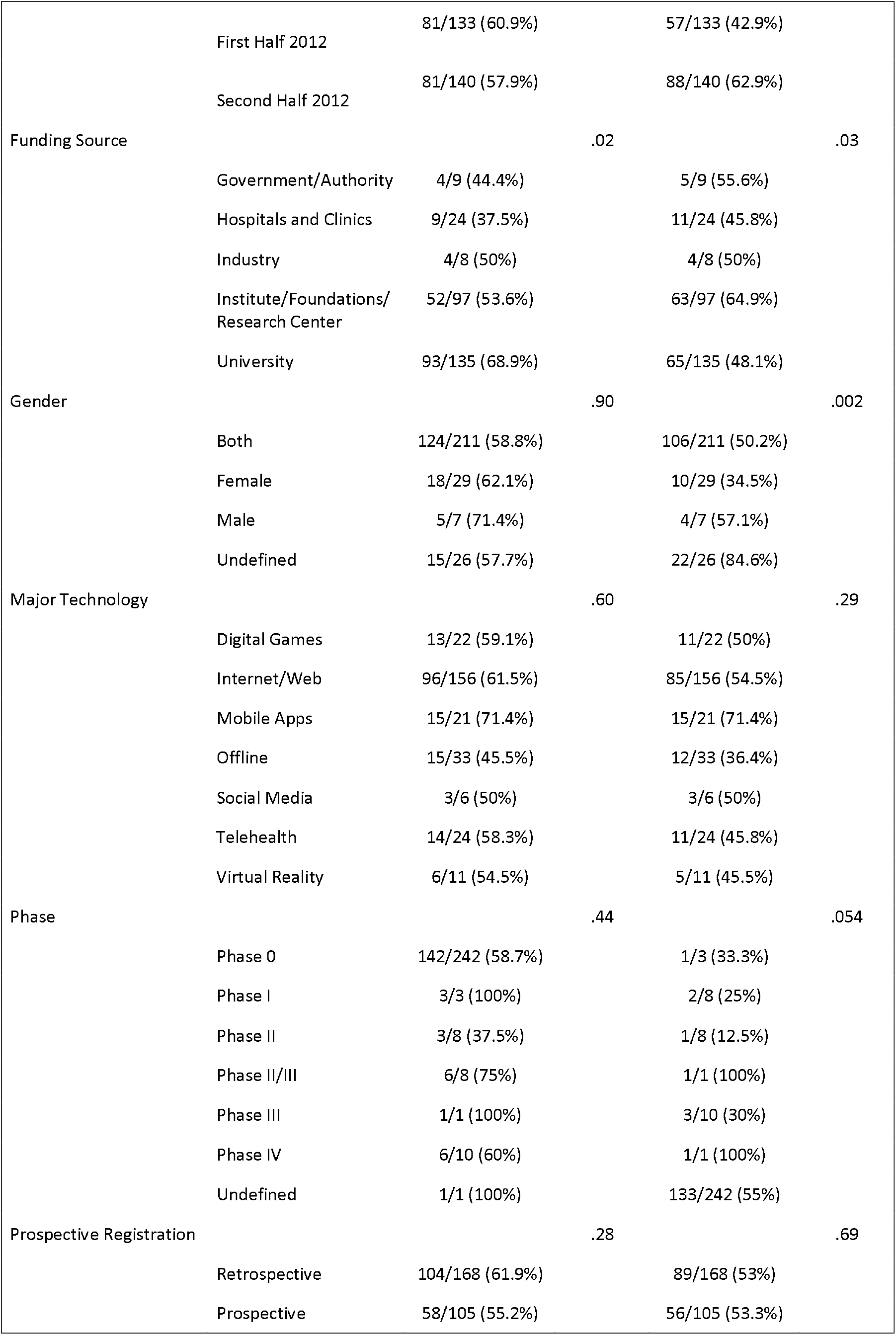
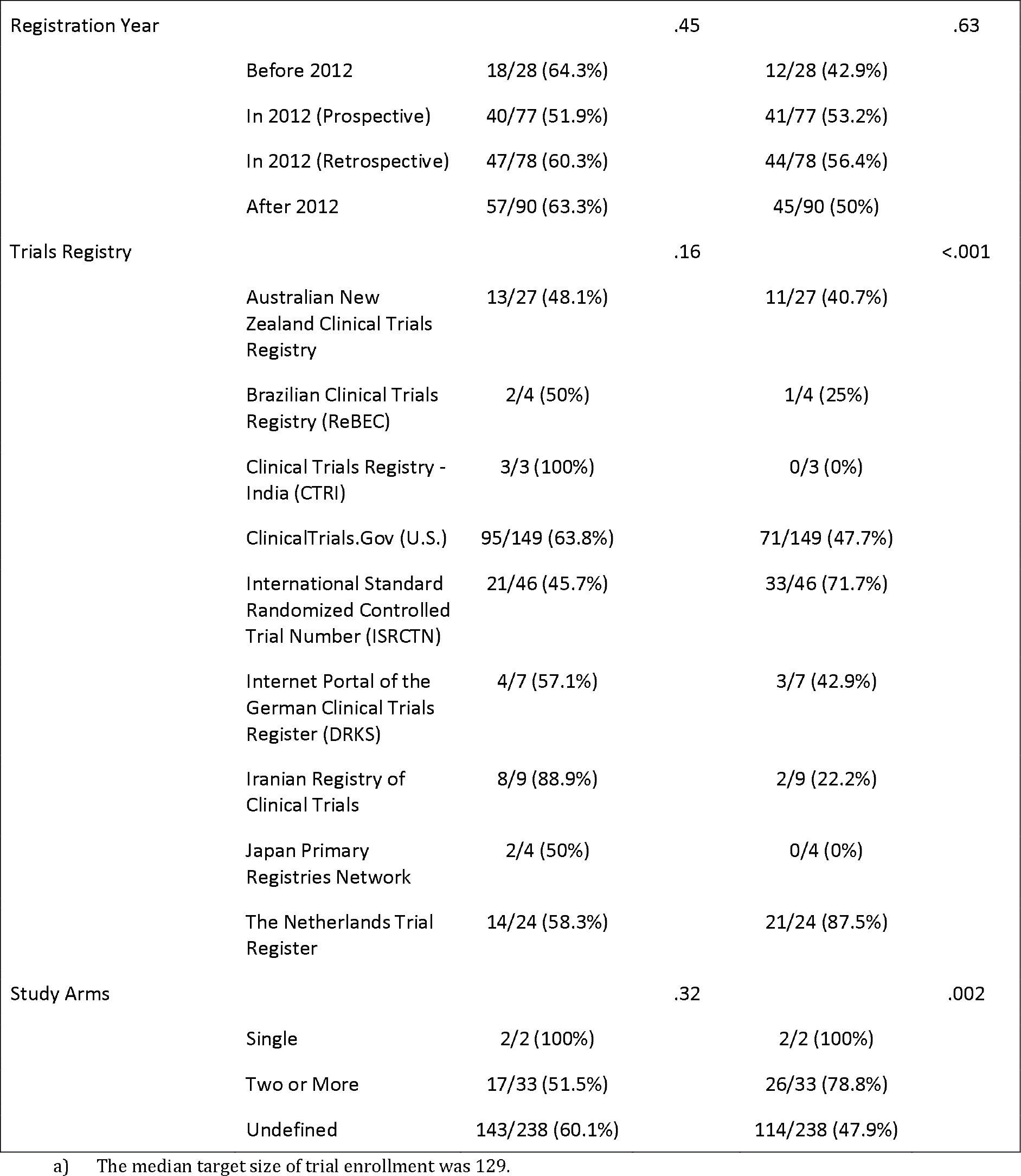
Recruitment Compliance and Inclusion of Trial Identification in Published Trials

The recruitment compliance rate was at 59.3% and nearly half of the trials 52% provided a reference to their respective trial identification number in their publications.

The Pearson Chi-square test results reported significant relationships (*P*<0.05) between trial recruitment compliance and trial funding source and trial size.

There were significant relationships (*P*<0.05) between trials with included the TRN and trial characteristics including trial age group, enrollment date, location, funding source, gender, trial size, trial registry, and study arms.

## Discussion

The primary objectives of this study were to examine the non-publication rate and prospective registration of digital health randomized clinical trials registered in any of the seventeen WHO ICTRP registries. A total of 417 randomized clinical trials met our inclusion criteria, were in forty different countries, and were registered in eleven trial registries.

We found that 34.5% of all included trials remain unpublished six years after the enrollment of trial participants. Nearly half of our included trials 56.1% were registered in the ClinicalTrials.gov registry, of which 36.3% remain unpublished. Our non-publication rate was higher compared to a 2018 study that reported 27% non-publication rate for digital health trials registered in ClinicalTrials.gov.[60] Although both studies focused on examining non-publication rates in digital health trials, their study design, data source and approach were quite different, hence a direct comparison would not be valid. The current study has a broader scope including 417 trials that were registered within one year (2012) in any of the seventeen WHO registries. The former 2018 study included 556 trials that were registered in the ClinicalTrials.gov registry only and were completed within three years, between 2010 and 2013.[60]

Our finding is also higher than that reported in another study with a non-publication rate at 29% for large randomized clinical trials, i.e. trials with at least 500 enrolled participants.[3] Other studies reported that nearly half of the included trials were not published, which is considerably higher than our own findings.[1,4,28] To explain these differences in non-publication rates, we postulate that the rapidly evolving technology elements of digital health trials could introduce extrinsic motivation to investigators to publish their results and stay ahead of the technology innovation curve.

Our results also showed that the vast majority 96.6% of digital health trials are funded by non-industry sponsors, such as universities, hospitals, medical institutes and research centers, that are more disciplined and obliged by scholarly ethics to publish their results. We speculate that industry sponsors would be more interested in the broader opportunity in the digital health marketplace beyond the realm of academia and best practices of randomized trials design.

We found statistically significant relationships between prospective trial registration and their funding sources. The lowest compliance with prospective trial registration was at 14.3% for industry-funded trials. In contrast to our present study, other studies assessing the prospective registration of clinical trials found that industry funded trials were more likely to be compliant with prospective trial registration compared to non-industry-funded trials.[61-70] However, we also found that only 14 (3.4%) trials of the included 417 trials were industry-funded which may limit the generalizability of our finding in comparison to other studies. For the 273 trials with identified publication in our study, the compliance in trial recruitment was at 59.3%, which is comparable to results from other studies reporting nearly one third of the clinical trials recruited their original target size.[71,72] Our Pearson Chi-square test reported a statically significant relationship between the compliance with trial recruitment and the trial size

(*P*=.009) and funding source (*P*=.02). Smaller trials were 1.3 times more compliant with recruitment than larger trials. We suspect that it would be easier to adequately recruit, manage and retain participants of smaller trials with a clear logistical, operational and feasibility advantages over larger trials. We also found that trials funded by university sponsors were 1.5 times more compliant with trial recruitment compared to trials funded by any other sponsors. This finding indicates that investigators of university sponsored trials are exceling at adopting best practices and strategies in trial design to improve participant recruitment, which is to be expected within the academic context of the university sponsors.

### Retrospective Registration of Digital Health Clinical Trials

Despite the emphasis on prospective trial registration introduced by the 2004 ICMJE mandate and the 2008 Declaration of Helsinki, we found that only 38.4% of all included trials were registered prospectively.[22,24] Similar findings were reported by two independent studies indicating that compliance with prospective trial registration was at 31%.[75,76] We hypothesize that investigators may be more inclined, or biased, to registering their trial only when submitting their results to peer-reviewed journals for publication.

### Selective Registration Bias of Digital Health Clinical Trials

We found a statistically significant relationship (*P*<.001) between retrospective registration and the registration-to-publication duration in digital health clinical trials, for which we coined a new term as “Selective Registration Bias”, or simply “Registration Bias”.

Within our sample of 273 published trials, the vast majority 95.7% of trials registered within one year before publication were registered retrospectively. Our results showed that investigators, who did not register their trials promptly prior to enrollment, were required or motivated to do so only when they submit their results to scholarly journals. There may be a number of contributing factors to this selective registration bias. Firstly, the investigator may have deferred the trial registration task until the completion of the trial and only when the results are finalized and ready to be published. Secondly, the investigator may also be not aware of the ethical expectation to register their trial promptly. Lastly, journal editors and peer-reviewers are likely to suggest registration of the submitted trial publication prior to accepting the publication submission. It’s important to establish and broaden the adoption of prompt trial registration requirement in ethical approval and guidelines of clinical trial design at the institutional and academic level.

### Challenges in Oncology Trials

We found statistically significant relationships between 5 different condition groups and the non-publication rate (*P*=.042) as well as compliance with prospective trial registration (*P*=.02). Our results indicate that investigators of oncology trials are the most compliant with prospective trial registration at 47.4% compared to investigators of trials of other conditions, but they seem to face more challenges to publish their oncology trial results with the highest non-publication rate reported in oncology trials at 63.2%. The significantly higher underreporting of oncology trial results suggests challenges in conducting oncology studies in the realm of digital health trials. These challenges may align with other studies indicating a number of barriers to conducting traditional oncology trials including eligibility, recruitment, follow-ups, and oncologist and patient attitudes.[62-64] However, we postulate that there may be a few other challenges that are specific to digital health oncology trials, particularly in the recruitment and enrollment stage of those trials. These explicit barriers may be explained by the treating oncologist, or patient, preferences to enroll in other non-digital health trials, such as experimental drug trials, with more measurable clinical outcomes. The results from our study validate this postulate through two distinct data points. First, we found only one oncology trial in our study that was funded by a pharmaceutical sponsor, indicating the lack of interest from pharmaceutical sponsors to invest in digital health trials. Second, we reported the lowest recruitment compliance rate for oncology trials at 42.9%, which is a clear indicator to enrollment barriers to digital health oncology trials.

### Compliance with Prospective Trial Registration in The Australian New Zealand Clinical Trials Registry (ANZCTR)

The Australian New Zealand Clinical Trials Registry (ANZCTR) leads with 89.7% compliance with prospective trial registration. Also, trials from Australia and New Zealand were leading with 89.5% compliance with prospective trial registration (the 0.2 percent decline is due to one trial that was registered in the ANZCTR registry and was not located in Australia or New Zealand). The high compliance with prospective trial registration in the ANZCTR registry was acknowledged by another study indicating an incremental trend in prospective trial registration from 48% to 63% for trials registered in the ANZCTR registry between 2006 and 2012.[61]

### Recruitment Compliance in the Middle East

Digital health trials in the Middle East had the lowest non-publication rate and the highest recruitment compliance at 26.7% and 81.1% respectively. These results were driven by Iranian trials registered in the Iranian Registry of Clinical Trials (IRCT). Compared to other WHO registries in our study, the IRCT trials had the lowest non-publication rate and the highest recruitment compliance at 25% and 88.9% respectively. Several studies reported to the legacy of the IRCT and its role in upholding best practices and ethical guidelines in the field of clinical research in Iran.[65-67] However, we found that prospective trial registration was the lowest at 8.3% for trials registered in the IRCT. Our results support findings from another study that reported on the rationale behind the low prospective trial registration rate at 8.3% for trials registered in the IRCT between 2008 and 2011.[68] All trials registered in the IRCT were funded by university sponsors. This finding aligns with another study that analyzed all trials registered in the IRCT until the end of 2015 and reported that 97% of those trials were funded by university and other governmental institutions.[52] The academic and public sponsorship may be a contributing factor to low non-publication rate and high compliance in recruitment of trials registered in the IRCT as they would likely encourage and promote a culture of adherence with trial design best practices and ethical guidelines. We also found that inclusion of the TRN in respective trial publications was the lowest for trials registered in the IRCT at 22.2%. Combined with the lowest prospective trial registration of those trials, we postulate that investigators of trials registered in the IRCT may be unaware of the ICMJE recommendation for prospective trial registration and the WHO re-affirmation to include the TRN in the trial publication to enable the easy linkage between the publication and registry entry of the respective trial.

### Recruitment Compliance in Low and Middle-Income Countries

We found significant differences (*P*=.002) in the relationship between recruitment compliance and trial location. We grouped Asia, the Middle East and the Americas without the U.S. as low and middle-income countries, and grouped high-income countries as Europe, U.S., Australia and New Zealand. The average compliance with trial recruitment in low and middle-income countries was at 51%, which is significantly higher than that in high-income countries at 30%. The significantly higher compliance in trial recruitment in low and middle-income countries may be explained by differences in clinical, regulatory, and economical standards in these countries, such as (1) access to a large population of potential trial participants (often with lower socioeconomic status and medical literacy), (2) lower cost of research resources, (3) less enforced, or developed, regulatory standards, and (4) if participation in a trial would provide access to, otherwise unavailable or unaffordable, medical care for the participants.[73,74,82] These differences raise ethical concerns in conducting clinical trials in these countries. These concerns are best described in article 20 of the Declaration of Helsinki emphasizing on the ethics in participant recruitment, and on the relevance and tangible benefits of the research outcomes to the participants population.[24,82] Local regulatory agencies, and ethics committees in low and middle-income countries need to be cautious about these ethical concerns and ensure the scientific integrity of digital health trials in their respective countries.

### Adherence to Best Practices in Clinical Trials from Europe

Europe was the region with the largest number of digital health trials in our study. European trials constituted 182(43.6%) of our included trials, and had the highest compliance with the inclusion of the TRN at 68.7%, and the second lowest non-publication rate at 28% tied with the Middle East. The overall lead in European trials is influenced by strong publication compliance demonstrated by the investigators of digital health trials in the Netherlands. The Netherlands National Trial Register (NTR) had the second lowest non-publication rate at 25%, and the highest rate in compliance with inclusion of the TRN in their respective trial publication at 87.5%. The latter finding was higher than another study of trials registered in the Netherlands National Trial Register that indicated that the compliance with reporting the TRN in respective trial publications was at 60%.[69]

### Time to Publication

The majority (88.3%) of the published trials in our study were published within five years after enrollment. Our finding is comparable to the results of a 2007 study indicating that trials with positive results were published within five years after enrollment and trials with negative results were published in six to eight years.[43] We postulated that small trials would be easier to conduct as they have fewer recruitment and enrollment challenges, hence they would likely be completed and published in a shorter time compared to large trials. To validate our postulate, we analyzed the relationship, and found statistically significant differences (*P*=.002), between trial size and enrollment-to-publication duration. We observed a reversed trend in publication between large and small trials at the fourth-year mark after trial enrollment. Small trials published their results 1.3 times more than large trials within the first four years after enrollment, after which large trials published their results 2.2 times more than small trials. This trend is likely driven by the longer amount of time required for the investigator of the larger trials to complete the enrollment and intervention for a larger group of participants.

### Limitations

Our study is the first to our knowledge that analyzed global digital health clinical trials registered in all the WHO recognized trial registries.[26] Our study did not consider any other registries that are not part of the seventeen WHO primary registries, such as the federal office of public health’s portal for human research in Switzerland, and the Philippine health research registry.[78,79] We acknowledge that not considering trials registries other than the WHO primary registries may have impacted the external validity of our study results.

Despite the ICMJE and WHO emphasis on trial registration, not all randomized trials are registered.[73,76] We did not include unregistered trials in our analysis, which may impact the internal validity of our study results.

We included trials that are registered and started the enrollment in 2012. We considered the enrollment and/or trial start date as provided in the registration information of the included trials. The registration information is provided manually and voluntarily by the registering investigators who are often overwhelmed with competing priorities and limited resources. Therefore, the enrollment date provided in the trial registries may not always be up-to-date or maintained promptly.

Lastly, our study only included registered trials enrolled in 2012. Our findings predate the recent 2015 WHO calls for improving public disclosure of trial results and the linkages between these results and the respective trial registry entries.[25]

## Conclusion

In the field of digital health randomized clinical trials, the adherence of investigators to the best practices of trial registration and result dissemination is still evolving. We analyzed digital health randomized clinical trials that were registered in the seventeen WHO recognized trials and started their enrollment in 2012. Within our included 417 trials, non-publication rate and retrospective trial registration were prevalent at 34.5% and 61.6% respectively. Our study indicated low compliance rate with recruitment and inclusion of the TRN in publication of the 273 trials, with identified publication, at 59.3% and 52% respectively. It would be advisable for the research community, from research ethics boards to journal editorial boards, to promote and advocate for better adherence to trial publication and registration. In particular, prospective trial registration could be mandated prior to obtaining institutional ethics approval and explicit reference to trial registration identification for all submitted trial manuscript for peer-reviewed journals could be enforced. Further research is required to identify contributing factors and mitigation strategies to low compliance rate with trial publication and prospective registration in digital health clinical trials.

## Methods

### Data Source

The WHO ICTRP is a free online portal that provides a unified access to trial registration information across different clinical trial registries.[26] As of September 2nd, 2018, the ICTRP database included 441,033 unique trial protocols from 17 different clinical trials registries. We utilized the advanced search feature in the ICTRP search portal to apply our search terms to the [Title] or [Intervention] field and downloaded the matching trials in XML format for further analysis.

### Statistical Analysis

We used the Pearson Chi-square statistic for bivariate analyses. All statistical analysis were performed in SPSS Statistics version 24 (IBM Corporation, Armonk, NY).

### Inclusion Criteria

We included all eHealth, mHealth, telehealth, and digital health related randomized clinical trials that are registered in any of the seventeen trial registries within the ICTRP database and include any information and communication technology component, such as:

- Online web and mobile application
- Internet, websites and personal computer application
- Digital games and social media application
- Telehealth and telemedicine components

We included trials irrespective of their recruitment status or trial phases. We limited our inclusion criteria to trials that started in 2012. We considered the enrollment date in the trial registries to indicate the start date of the included trials.

### Enrollment Date Justification

We aimed to allow for longer publication cycles to account for late publications of included trials. We reviewed existing studies that reported that investigators of clinical trials may take up to three years to complete their trials and another two to three years before they would publish their results.[27,40-42] A 2007 study indicated that clinical trials with positive results were published four to five years after their start date whereas trials with negative results would take six to eight years to publish their results.[43] We informed our design by this evidence and chose to include trials that started their enrollment in 2012, i.e. six years prior to conducting our research in September, 2018. We limited the scope of our analysis to one year only, i.e. 2012, to keep our study manageable and feasible.

### Exclusion Criteria

We excluded registered clinical trials that were not randomized or did not include any digital components in their intervention. Trials that merely utilized short messages service (SMS), phone-calls, emails or video communication without any other interactive components were also excluded. We excluded trials that only reported on computerized or online surveys and questionnaires.

### Search Terms

We developed a comprehensive list of search terms and phrases through an iterative process as explained in Appendix I - Determination of Search Terms and Phrases. Our final set of 86 search terms included:

“smartphone,” “smart-phone,” “cellphone,” “cell-phone,” “cellular phone,” “cellular-phone,” “cell phone,” “mobile phone,” “health application,” “mobile application,” “phone application,” “touch application,” “health app,” “mobile app,” “phone app,” “touch app,” “multimedia,” “multi-media,” “multi media,” “text reminder,” “short message,” “text message,” “messaging,” “texting,” “sms,” “email,” “e-mail,” “electronic mail,” “iphone,” “android,” “ipad,” “online,” “on-line,” “e-Health,” “eHealth,” “mhealth,” “m-health,” “internet,” “etherapy,” “e-therapy,” “e-therapies,” “information technology,” “communication technology,” “information application,” “electronic application,” “well-being application,” “informatic,” “computer,” “digital,” “Web,” “telehealth,” “tele-health,” “tele-monitoring,” “telemonitoring,” “tele-medicine,” “telemedicine,” “tele-rehabilitation,” “telerehabilitation,” “tele-consult,” “video consult,” “video-consult,” “video conferenc,” “video-conferenc,” “skype,” “social media,” “social-media,” “social network,” “social-network,” “social app,” “facebook,” “twitter,” “tweet,” “whatsapp,” “wearable,” “fitbit,” “wii,” “nintendo,” “kinect,” “xbox,” “playstation,” “game,” “gamif,” “gaming,” “hololens,” “virtual reality,” “augmented reality,” “google glass.”

### Data Extraction

We downloaded the XML files for the 417 matching clinical trials in the ICTRP search portal. We transformed these files into a tabular format and imported them into a local SQL Server Database for further data preparation and analysis.

### Conditions

The content of the condition field in the ICTRP dataset is a free-text description of the trial conditions. We were able to consolidate a total of 375 unique condition descriptions of the 417 included trials to 5 distinct condition groups as described in Appendix II - Classification of Trials Condition Groups.

### Prospective Trial Registration

The data export from the ICTRP dataset did include a field to indicate whether a trial was registered retrospectively.[44,45] However, our analysis showed that this field did not include correct information for a substantial number of trials within our sample of 417 included trials. We therefore chose to evaluate prospective trial registration based on the actual difference between the registration and enrollment dates as described in Appendix III - Identification of Prospective Trial Registration.

### Primary Sponsors

We analyzed the primary sponsor field in the data extract from the ICTRP dataset.[44] Within our 417 included trials, we categorized the primary sponsors as 205 “University”, 137 as “Institute/Foundation/Research Center”, and grouped all the remaining 75 sponsors under “Others” as they had minimal representation within our included dataset. The remaining 75 sponsors, that we categorized as “Others”, included 14 industry, 18 government, and 44 hospital or clinic sponsors.

### Major Technology

We evaluated the digital components utilized within the included trials and provided a classification for major technology used within the respective interventions. Details of our classification approach are provided in Appendix IV – Classifications of Trials Major Technology.

### Identification of Randomized Trials

We were able to identify randomized trials through a text match search in the following three fields in the data export from the ICTRP dataset: “Study_design,” “Public_title,” and “Scientific_title”. We searched for matching words that included the term ‘Random’ and did not include the term ‘Non-Random’.

### Identification of Publication

We identified existing publications through an automated and a manual publication identification process. The automated identification process included a PubMed search by every trial registration ID as well as a review of listed publication references and citations for trials registered in the ClinicalTrials.gov registry. The manual process included a pragmatic search in PubMed and Google based on a combination of search terms concatenated information from trial titles, investigators, location/city and institution. We only considered trial publications that reported at least one of the primary outcome measures of the underlying trials. Complete details of the publication identification processes are described in Appendix V – Identification of Trials Publication.

## Data Availability

The data source for this research was the publicly available trial registration information in the the WHO International Clinical Trials Registry Platform (ICTRP). The data set for the reported results of this research is available upon request.

## Data availability

Clinical trials registration data and material were downloaded from the WHO ICTRP database or directly from the website of the primary registries in the WHO registry network. The seventeen clinical trials registries included in this study are publicly available online: https://www.who.int/ictrp/network/primary/en/

## Code availability

Computer code used in this study is available upon reasonable request to the corresponding author and under a collaboration agreement.

## Acknowledgements

None declared.

## Abbreviations

ANZCTR: Australian New Zealand Clinical Trials Registry
ICD: International Classification of Diseases
ICMJE: International Committee of Medical Journal Editors
ICTRP: International Clinical Trials Registry Platform
JPRN: Japan Primary Registries Network
IRCT: Iranian Registry of Clinical Trials
ISRCTN: International Standard Randomized Controlled Trial Number
NIH: National Institutes of Health
NLM: United States National Library of Medicine
MeSH: Medical Subject Headings
PACTR: Pan African Clinical Trial Registry
WHO: World Health Organization
XML: eXtensible Markup Language

